# CorCast: A Distributed Architecture for Bayesian Epidemic Nowcasting and its Application to District-Level SARS-CoV-2 Infection Numbers in Germany

**DOI:** 10.1101/2021.06.02.21258209

**Authors:** Anna-Katharina Hildebrandt, Konstantin Bob, David Teschner, Thomas Kemmer, Jennifer Leclaire, Bertil Schmidt, Andreas Hildebrandt

## Abstract

Timely information on current infection numbers during an epidemic is of crucial importance for decision makers in politics, medicine, and businesses. As information about local infection risk can guide public policy as well as individual behavior, such as the wearing of personal protective equipment or voluntary social distancing, statistical models providing such insights should be transparent and reproducible as well as accurate. Fulfilling these requirements is drastically complicated by the large amounts of data generated during exponential growth of infection numbers, and by the complexity of common inference pipelines. Here, we present CorCast – a stable and scalable distributed architecture for the reproducible estimation of nowcasts suitable for pandemic scenarios – and its application to the inference of district-level SARS-CoV-2 infection numbers in Germany.

## 1 Introduction

The SARS-CoV-2 pandemic that emerged in late 2019 has clearly shown the need for accurate, timely, and fine-grained infection statistics. To assess infection risks for different parts of the population, e.g., different age groups, the current number of daily infections for that sub-population in the geographic region of interest is obviously crucial. Corresponding datasets also allow for the estimation of the current replication number R, and thus enable a judgement about the efficiency of current anti-epidemic measures, such as social distancing or mandatory wearing of private protective equipment such as face masks.

Unfortunately, the data required for these applications is typically not directly available due to a number of problems of both fundamental and practical nature, some of which are exacerbated by the particular properties of SARS-CoV-2. To illustrate these problems, let us assume that a person *p*_*i*_ is infected at date *x*_*i*_. After an incubation period, the disease begins at date *s*_*i*_, is diagnosed at date *d*_*i*_, and finally registered in a database at date *r*_*i*_, together with relevant information about *p*_*i*_ (such as gender, age, and geographical region). The whole process is summarized in Figure 1.

**Figure 1:**
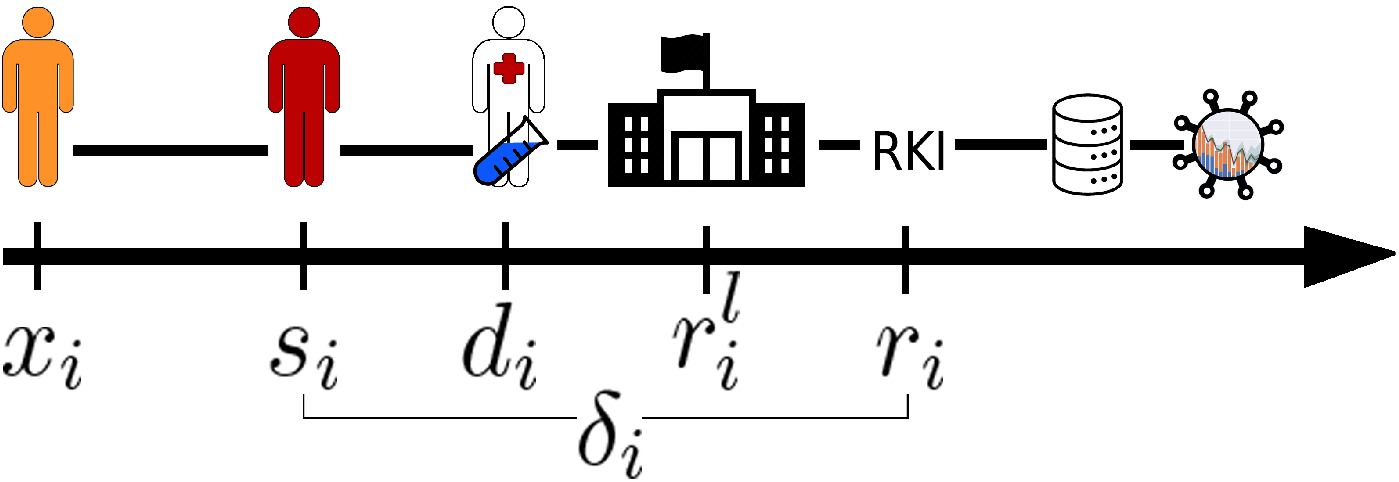
Timeline of dates during infection and reporting. A person gets infected at date *x*_*i*_, the disease begins at date *s*_*i*_, is diagnosed at date *d*_*i*_ and gets registered at dates 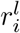 at the local health authority and *r*_*i*_ at the RKI, Germany’s public health institute. *δ*_*i*_:= *r*_*i*_− *s*_*i*_ is called the reporting delay. After registration at RKI, the case is reported in a database and can be fetched by CorCast.

Obviously, not all infections are identified. In fact, it is generally unknown, at least until detailed antibody studies can be performed, what the true infection rate is. This is particularly true for diseases such as SARS-CoV-2, which lead to a large percentage of asymptomatic or only mildly symptomatic cases. A further complication arises from the fact that, even for identified cases, the date of disease onset *s*_*i*_ is often unknown, for instance in asymptomatic cases that were identified through contact tracing or routine mass testing. In particular, it is not generally possible to assume that *s*_*i*_ = *d*_*i*_, and in particular that *s*_*i*_ = *r*_*i*_. Instead, each case has an individual *reporting delay* that is composed of the delay between disease onset and diagnosis as well as the delay between diagnosis and reporting, and this delay is unknown for a large percentage of the reported cases.

Owing to the reporting delay, information about infection numbers at the most recent days is necessarily inaccurate, or rather, incomplete. The goal of *nowcasting* of infection numbers is thus to infer an estimate of the true infection numbers based on the current, incomplete counts and the reported infection history [16]. This task is often decomposed into two phases [13]: the imputation of disease onsets for cases that have already been reported, but for which the true onset is unknown, and the nowcasting based on this data. These phases consist of a number of steps, including data ingestion, data cleanup and preprocessing, imputation of disease onset, nowcasting, postprocessing, validation, and reporting.

### 1.1 Our contribution

When developing a Bayesian model for imputation and nowcasting of the SARS-CoV-2 infection numbers in Germany at a district level, we encountered a number of challenges. For instance, each step or module should be individually exchangeable, which greatly simplifies development and debugging and allows for testing and validation of combinations of module variants. To ensure reproducibility, it is imperative that data, modules, and workflows can all be versioned in a manner that allows exact recreation of the conditions at a specific point in time. Due to the large amounts of data generated in the exponential phase of a pandemic, pipelines further need to scale out sufficiently. Finally, deployment and maintenance of the system needs to be addressed.

Despite their crucial importance, these practical steps of implementation, deployment, operation, and maintenance are often ignored in the literature.

Here, we report on a stable and scalable distributed system, called CorCast, for the reproducible estimation of nowcasts suitable for pandemic scenarios. CorCast’s implementation is highly modular – indeed, retargeting CorCast to other geographical regions or other diseases can be achieved by adapting only the data ingest and dashboarding modules. Similarly, the influence of different preprocessing methods, different imputation models, or different now-cast models on the final nowcast can be systematically studied. By basing CorCast on the Pachyderm framework [22], we guarantee reproducibility. Scaling out is achieved through the use of big data concepts and a scalable Kubernetes deployment.

To validate our architecture, we build a full pipeline for the nowcasting of Covid-19 infections in Germany on a district, state, and country-wide level (cf. Figure 2).

**Figure 2:**
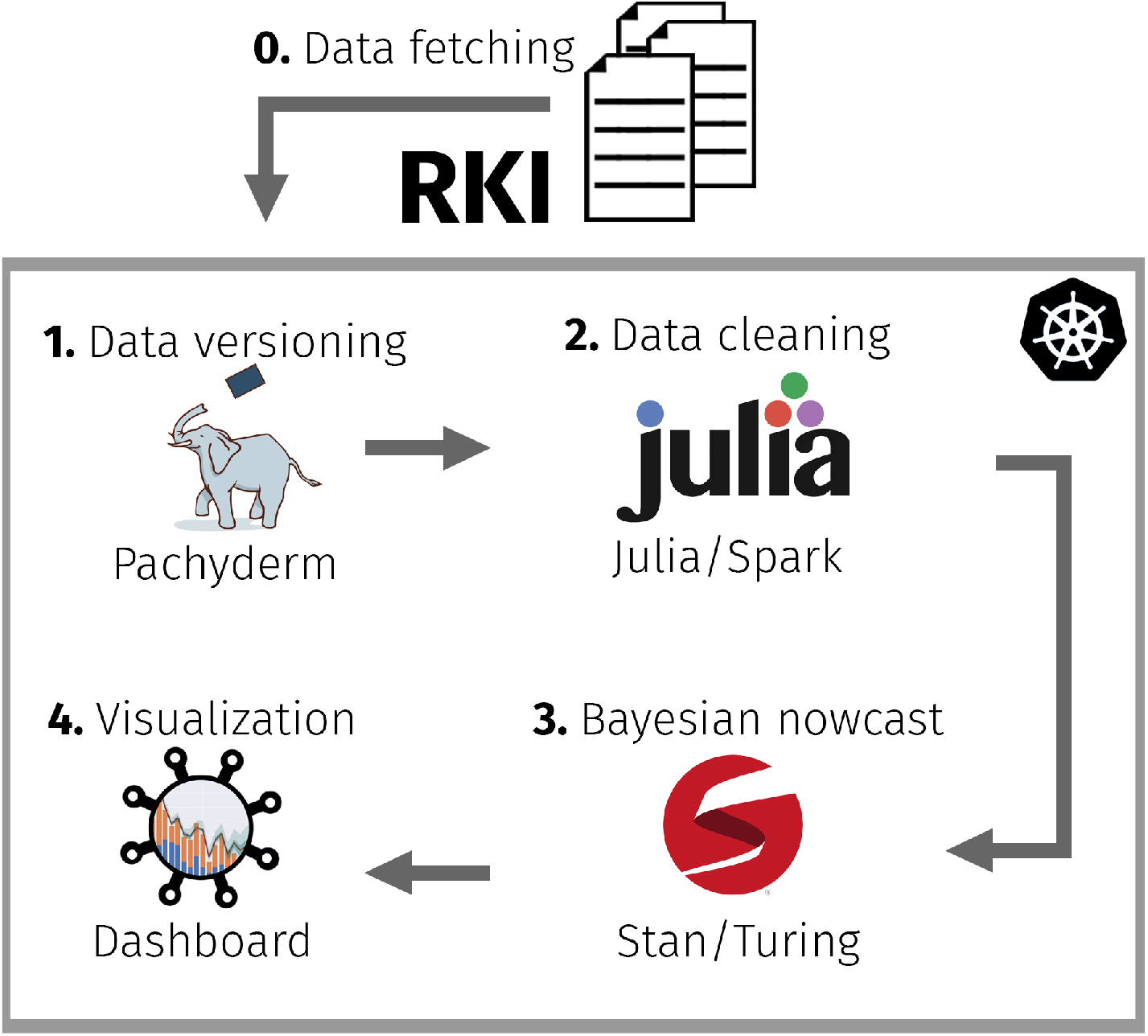
Graphical representation of the nowcast workflow. Data is fetched from the Robert Koch Institute, Germany’s public health institute, once per day (0). All versions are saved and checked in for tracking with Pachyderm (1). Data needs to be cleaned and standardized by a set of custom scripts written in the Julia programming language (2). Afterwards, imputation and nowcast can be performed (3). Lastly, processed data and nowcast results are visualized via a custom designed dashboard (4). All CorCast components are orchestrated through Kubernetes.

### 1.2 Related work

Related work, such as [16] followed, e.g., by [28] and [13], mainly focused on statistical modeling for nowcasting, while [7] and [1] dealt also with forecasting. Software packages for disease monitoring are covered, e.g., in [25], [26], and [15], but to the best of our knowledge, no general framework for the development of such prediction techniques has been proposed prior to this work.

## 2 Results

The main result of this work is the CorCast system as a computational pipeline. In the following, we will describe its essential components and the application to Germany as a proof of concept.

Our proposed architecture is sketched schematically in Figure 2. Inference is based on probabilistic programming using Stan [5] and Turing [9]. All system services are deployed on a Kubernetes cluster [2] for standardized deployment and operations, including simple horizontal scaling on demand. On the cluster, a Pachyderm installation [22] orchestrates compute pipelines and datasets. Pachyderm also keeps track of revisions of individual compute modules and pipelines as well as of input, intermediate, and result datasets. Finally, a Flask- [23] and Dash-based [24] web service, deployed on the Pachyderm cluster as a service, provides a graphical interface to the ingested and processed data as well as to the imputation and nowcast results.

Owing to the high case numbers of the pandemic, each individual module needs to be highly efficient. At the same time, to facilitate extensions and improvements of the pipelines, the module implementations should be intuitively readable and high-level to hide many implementation details required for scaling to modern compute hardware. We achieve this goal through the use of the Julia language [3].

In the following, we will discuss the individual components of this system in more detail.

### 2.1 Data ingest and preprocessing

The first step in our pipeline is the collection of relevant infection data. In Germany, for instance, infection data is collected locally at the health departments at the district level (a political organizational unit below the state). Those departments collect the relevant information and further transmit it to the Robert Koch Institute (RKI), Germany’s public health institute. At the RKI, collected information is processed to ensure compliance with privacy regulations, and then published daily. The format of this publication is rather unusual and cumbersome. In addition, the reported dates of interest (infection date, reporting date, and dates of recovery or death) cannot, in general, be uniquely assigned to individual cases^1^. Expanding on our notation from Section 1, let us assume that an individual *p*_*i*_ contracts the disease at date *x*_*i*_ and is diagnosed at date *d*_*i*_. This diagnosis is reported locally, i.e., at district level, at date 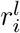 and transmitted to the RKI, which receives the case information at date^2^ *r*_*i*_. Each day, the RKI then publishes a spreadsheet containing information on all previously received cases.

Extracting the required information from these data publications is a challenging task in itself. For more details, c.f. Sec. 4.1.

### 2.2 Imputation of disease onset

The goal of this stage is to impute missing disease onset information. Let *C* denote the set of all cases, *C* ⊇ *K* := {*i* ∈ *C*|*s*_*i*_ is known} the set of cases with known disease onset, and *C* ⊃ *U* := *C* \ *K* the set of cases with unknown onset. Our task is now to infer *s*_*i*_ ∀_*i*_ ∈ *U* based on the information contained in *K*. To this end, we build Bayesian (hierarchical) models for the *reporting delay δ*_*i*_:= *r*_*i*_− *s*_*i*_. Inference is then achieved by sampling *δ*_*i*_ from the posterior for each case in *U* and using *δ*_*i*_ to finally predict^3^ *s*_*i*_ = *r*_*i*_− *δ*_*i*_.

In our framework, we implement several different statistical models for the delay distribution in two probabilistic programming frameworks (Stan [5] and Turing [9]). The baseline is a trivial model that does not take any covariates of interest into account, see Section 4.3 for details. However, even the baseline model fits the data remarkably well. Figure 3 shows an example.

**Figure 3:**
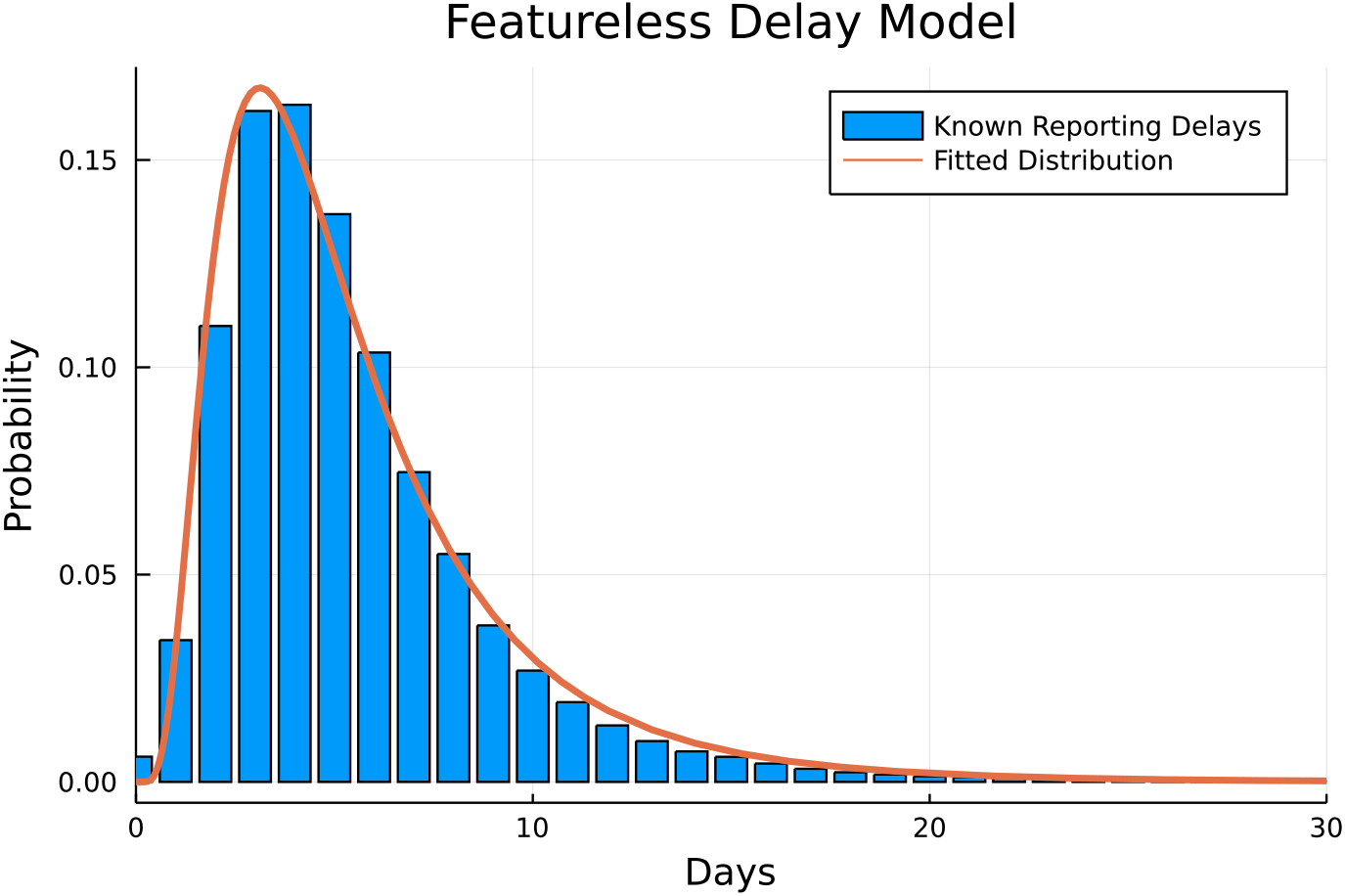
The delay histogram (delays shifted by 1 day to prevent values of zero) against a LogNormal distribution with parameters generated from the mean of 4 MCMC chains with 2,000 samples each, generated by the NUTS [17] sampler fed with a subset of 1,918,308 known delays.

In a more complex model we included additional covariates, such as age group, gender, or district of the cases. Since it is reasonable to assume that reporting delays might change over time (e.g., owing to varying test rates or overloads of institutions during high-incidence intervals), the more sophisticated model also includes a slowly varying time-dependent term, e.g., modelled by a spline.

Additionally, the data has an obvious hierarchical component – in Germany, districts are located within states. A multi-stage hierarchical model thus seems appropriate to represent dependencies by partial pooling: the means and variances of the delay distribution in each district should ideally be able to learn from the means and variances of each state, and those of the states from those of the country as a whole.

Figure 4 shows a graphical representation of the model. See Section 4.3 for a detailed description of the model.

**Figure 4:**
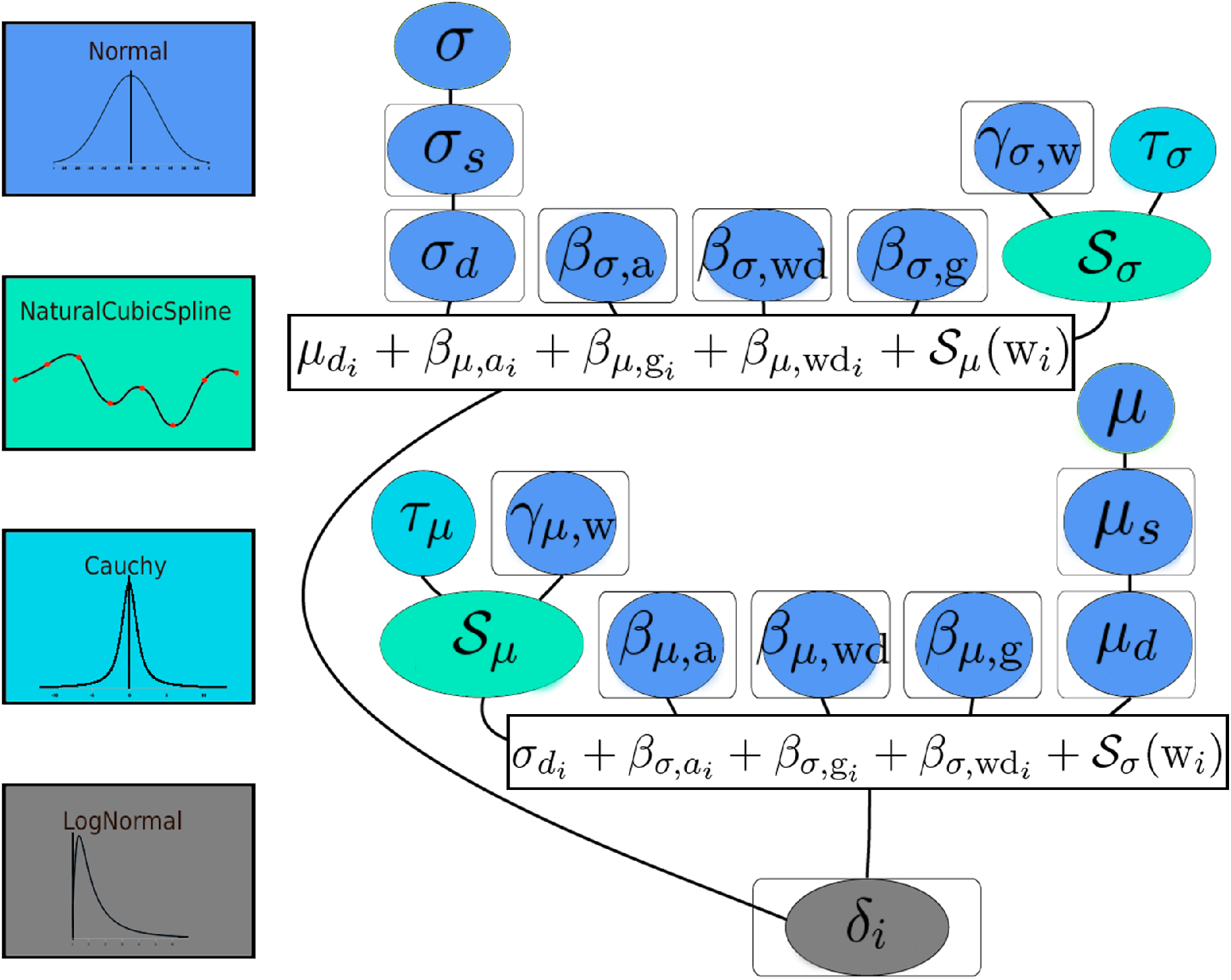
Imputation model with covariates age group, gender, district, weekday, and number of weeks since the start of the pandemic. Ellipses denote random variables, ellipses inside boxes indicate multidimensional variables, the terms in boxes indicate dependent random variables and lines indicate dependencies. The prior distributions used are encoded by color. Note the hierarchical structure of the spatial covariate. Due to computational constraints the model was evaluated in several steps. Details are given in Section 4.3.

We implemented our models in both Stan [5] (using CmdStan.jl [6]) and Turing.jl [9] and draw samples from the posterior using the NUTS [17] sampler to determine probability distributions for the model parameters. From these distributions, we can finally impute infection dates by drawing – for each case with unknown disease onset – delay values from the resulting posterior predictive distribution, conditioned on the appropriate covariates (e.g., the age group, gender, or district associated with the case).

Here it is worth noting that the delay distributions (cf. Figure 3) tend to have a rather pronounced peak, typically close to 7 days. If we would sample delay values multiple times for each case with unknown disease onset to average over the posterior predictive distribution, we would thus almost always assign delays of approx. 7 days. Instead, delay values are drawn once per case and used to compute disease onsets. We then group by disease onset and aggregate cases. Repeating this process multiple times allows to compute confidence intervals for the number of new infections per day.

### 2.3 Nowcasting infection numbers

During an epidemic, infection numbers cannot be observed in real time. Due to the reporting delay between the date *x*_*i*_ of disease onset and *r*_*i*_ of receiving the case information at the central case registry (the RKI in Germany), information about current case numbers will necessarily be incomplete. If we assume that the reporting delay *δ* is bounded by a reasonable maximum *D*, we can expect, at each day during the epidemic, to be notified about additional cases with disease onset in the last *D* days (we assume that reporting delays are generally *>* 1).

We adopted a model from [13], which in turn extends earlier work by [16]. The details are given in Section 4.4.

To provide nowcasts at the different levels of the hierarchy (district, state, and country), we group the data by district and run NUTS-sampling on each of the resulting 412 datasets (one for each district), followed by one run for each of the 16 states and one for the whole of the country. This process is quite time consuming and takes roughly 5 hours on three AMD EPYC™ 2nd Gen. worker nodes with 8 cores and 32 GiB RAM each.

### 2.4 Evaluation of nowcast

The evaluation of nowcast results is not a trivial task. In Section 4.5, we summarize the relevant literature. CorCast contains an evaluation module that implements the WIS-score described in that section.

### 2.5 Dashboard functionality

CorCast provides a dashboard feature through Pachyderm’s service pipeline mechanism. The dashboard is implemented in Flask [23] and Plotly’s dash framework [24], and can be easily extended and adapted. The dashboard for the CorCast model of district-level imputation and nowcasting in Germany can be found at https://dashboard.covid19.hildebrandtlab.org/ (cf. Figure 5).

**Figure 5:**
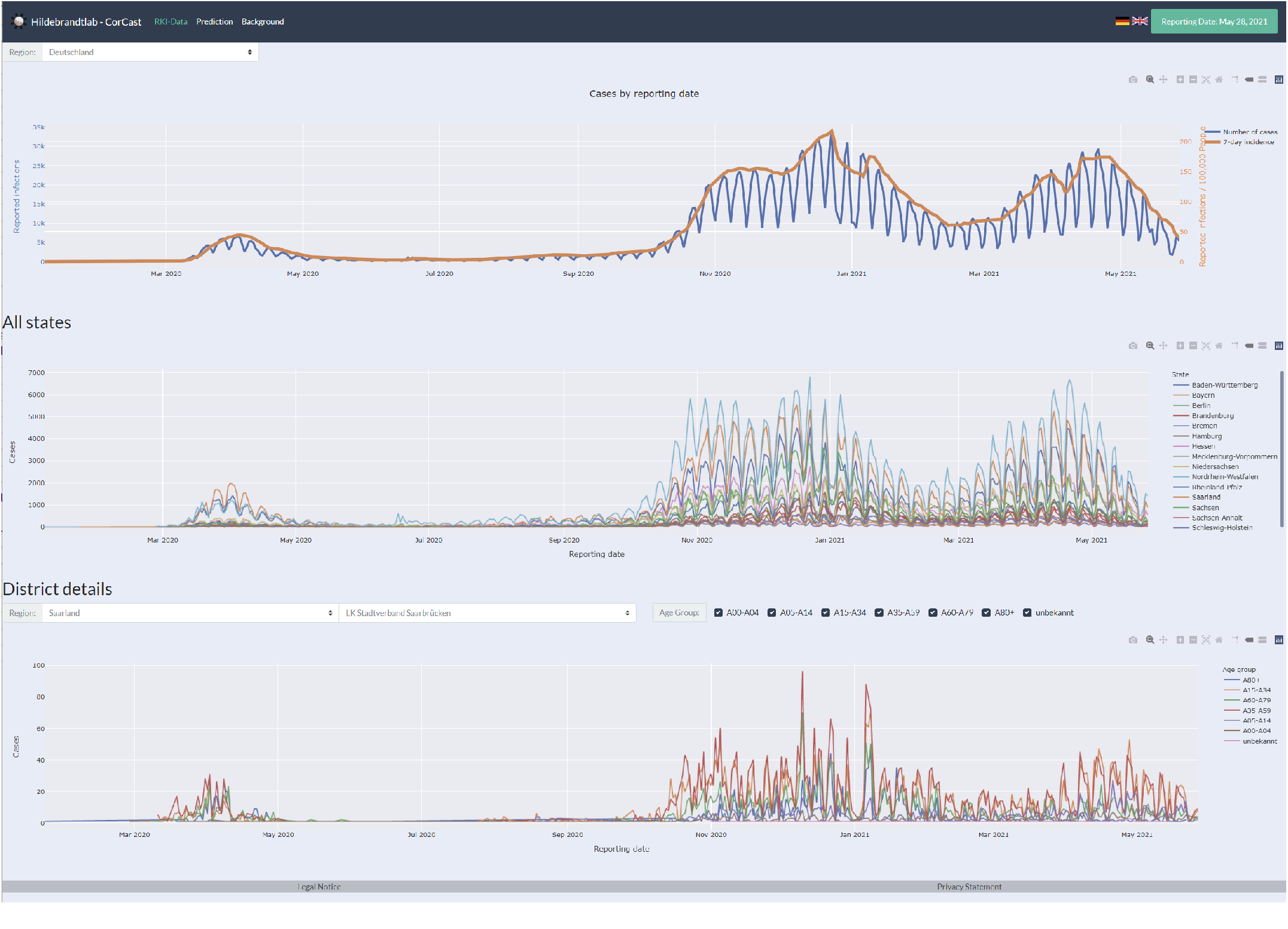
The CorCast dashboard.

## 3 Discussion

Several advantages distinguish the CorCast system described in this work. Here, we want to summarize the most important of these, namely (i) reproducibility (ii) modularity and extensibility (iii) scalability and (iv) standardized deployment and operations.

### 3.1 Reproducibility

Reproducibility is a cornerstone of science that can be very challenging to achieve. In complex computational systems, it is typically insufficient to merely guarantee accessibility to input data sets and informal descriptions of the employed programs and workflows. Modern scientific computer programs often rely on a large number of third-party dependencies, such as support libraries. Even small variations in the computational infrastructure can lead to differing outcomes, either through the introduction of bugs [4] or through numerical effects that lead, e.g., to convergence to different minima. True reproducibility hence requires reliable versioning of data, programs, and workflows. CorCast achieves this goal by relying on three key technologies: Git, for versioning source code, Docker containers [20] for versioning applications, and Pachyderm [22] for versioning data and workflows.

A side effect of incorporating reproducibility tightly into the system design is a greatly simplified evaluation and comparison of different steps in the nowcasting pipeline. Imagine, for instance, that we want to compare a novel Bayesian model for imputation of disease onset with respect to its influence on the final nowcast. Through the versioning of data and workflows, we can recreate the exact information used to perform the nowcasts at any former date since the initial data commit of the CorCast system, exchange the imputation module through our new candidate, run the full nowcasting pipeline, and finally evaluate the performance and compare it to the historic performance of our former model, which we can again query from the corresponding data repository.

### 3.2 Modularity and Extensibility

CorCast has been designed as a highly modular pipeline, with components responsible for ingesting data, processing and cleaning of the raw inputs, imputation of disease onsets, nowcasting, evaluation, and interactive visualization. Each of these components can be easily replaced, but it is also a simple task to add steps to the pipeline, or to provide alternative implementations for each step in the pipeline (such as competing nowcasting models) and to run them in parallel. To give an example of the implications of this extensibility, we want to stress that the current implementation of CorCast is focused on nowcasting SARS-CoV-2 infection numbers in Germany, but that it would be a simple matter of extending or replacing the ingest nodes of the pipeline to extend it to other countries, or to adapt it for different infectious diseases.

### 3.3 Scalability

While it might not be obvious at first glance, the data generated during a pandemic grows very rapidly. Merely reporting summary statistics, i.e., the number of new infections per day, is typically insufficient, as the progression of the epidemic can vary significantly in different sub-populations (differentiated, e.g., by age, gender, or location). To account for these differences, all differentiating features (including the date of disease onset) will have to be reported. In practice, much of the reporting hence happens at the level of individual cases, which grows rapidly during the pandemic phases of exponential growth. Further complications arise through inconsistencies and retrospective modifications that have to be treated carefully in the data preparation stage. In the case of German infection numbers as provided by the Robert Koch Institute, this requires iterating over all previous case reports, where each case report contains information about every single case since the start of the pandemic, leading to quadratic effort. At the time of writing, the uncompressed data size for this correction step is approximately 43.4 GiB. The large case numbers also render Bayesian inference rather expensive. At the time of writing, imputation of disease onset takes roughly 5 hours, nowcasting roughly 4 hours, on a Kubernetes cluster with one dedicated master and three AMD EPYC™ 2nd Gen. worker nodes with 8 cores and 32GiB RAM each. To handle the large amounts of data and the computational effort, we have taken care to expose opportunities for parallelism, e.g., by separating inference models by geographic region, by running multiple shorter MCMC chains in parallel, or by vectorization- and parallelization primitives inside the computational nodes. Scheduling of parallel computation is achieved through Pachyderm’s parallelization primitives. Without these measures, it would be essentially infeasible to produce daily nowcast results at our level of granularity.

### 3.4 Standardized Deployment and Operations

CorCast uses DevOps principles to facilitate deployment and operations. Following an infrastructure-as-code (IaC) approach, all components are provisioned and managed using Terraform [14] and Helm [8] files. All compute nodes are realized as Docker containers and all workflows encoded as Pachyderm pipelines in JSON format. Orchestration and managing of system resources (monitoring, up- and down-scaling, upgrade handling, etc.) are realized through Kubernetes and Prometheus. As a result of the IaC approach, all information required to set up a CorCast cluster is under version control in the CorCast Git repositories. Scaling the existing instance or setting up a new instance on a variety of cloud providers or on premise is trivial. Hence, CorCast can be easily adapted and deployed by researchers interested in epidemics now- or forecasting.

### 3.5 Summary

CorCast is a highly flexible and scalable framework for the implementation of computational statistics and machine learning approaches to epidemiology that provides automatic reproducibility. In addition, the CorCast instance described in this work is, to the best of our knowledge, the only currently available implementation for nowcasting of German Sars-CoV-2 infection numbers that can provide daily updates for disease onset imputation and nowcasts on a district level. In future work, we intend to add additional models to the CorCast system, such as the stable inference of replication numbers, and want to develop pipelines for other geographic regions.

## 4 Methods

### 4.1 Preprocessing

The RKI provides so-called “publications” that contain sufficient information to query the number of infections, recoveries, and deaths as a function of 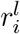, i.e., the date at which they were recorded locally at the health department of the district. Unfortunately, the files do not contain the date at which they were recorded centrally by the RKI, *r*_*i*_. Since much of the information provided by the RKI, such as imputed disease onsets and nowcasts, seems to be predicted based on *r*_*i*_ rather than on 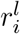, the lack of this information is rather unfortunate. To recreate *r*_*i*_, it is necessary to keep an archive of all previous daily data publications of the RKI (a task for which Pachyderm is ideally suited), and iterate backwards to identify at what date a case first appeared in the RKI publications. Since the data publications also contain corrections, which are modelled as deletions of cases in previous publications and new insertions of these cases, recreating *r*_*i*_ is non-trivial and error-prone. In addition, the publications contain numerous inconsistencies and errors that have to be identified and corrected for as accurately as possible. Finally, the process is made considerably more cumbersome by the fact that the data format varies over time. Dates, in particular, are stored in at least four different formats, depending on the date of the publication and the data column. Hence, parsing and preprocessing the input data is relatively complex. At the same time, the preprocessing needs to be highly efficient: individual cases have to be tracked through hundreds of files, each of which needs several steps of data conversion, processing, and quality control. Coupled with the large case numbers of the pandemic, preprocessing becomes quite resource intensive.

The ingestion pipelines at the start of our Pachyderm workflows start with a module that downloads the current RKI data publication and archives it in a Pachyderm repository. In the next step, the newest publication is processed and converted into a dataframe, which is then serialized to another Pachyderm repository. Simultaneously, we attempt to reconstruct the missing *r*_*i*_ values through iteration over all previous data publications^4^. The results are again stored in a Pachyderm repository. This approach allows us to base further processing steps on either the RKI-provided 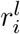 or on the missing *r*_*i*_ values.

### 4.2 Bayesian hierarchical modeling

Bayesian modeling is statistical modeling that is centered around Bayes’ theorem^5^: One starts with formulating possible hypotheses and assigns probabilities (prior probabilities *p*(*H*)) to them before comparison to the data *D* and then computes the conditional probabilities of each hypothesis given the data (posterior probabilities *p*(*H*|*D*)).

This approach has the advantage that already available information can be leveraged in a transparent way and the uncertainty of estimated quantities can be quantified as well. Furthermore, the robustness of predictions can be greatly improved by using posterior weighted predictions.

When dealing with data that arises in systems with a hierarchical nature, i.e., follow some tree-like structure, a common technique is to split the prior probability distributions into a product that resembles this structure, giving rise to hierarchical models [10].

Exact inference of Bayesian hierarchical models is hindered by fact that the integral d*H* ∫*p*(*D*|*H*)*p*(*H*), where *p*(*D*|*H*) denotes the likelihood, is typically intractable by symbolic expressions. Thus, numerical methods are used in practice, which are mostly sampling-based. These Markov chain Monte Carlo (MCMC) methods generate samples that are distributed according to the posterior and thus allow to compute approximations to quantities of interest on that sample.

While the classical Metropolis-Hastings algorithm [21] provides asymptotically correct samples, more advanced algorithms like [17] from the family of Hamiltonian Monte Carlo algorithms yield a better performance in terms of required sample size and correlation of samples. As Hamiltonian Monte Carlo requires the computation of gradients, several software packages, e.g., [5], [9], and [27], were developed to leverage automatic differentiation of model expressions.

### 4.3 Imputation models

Our baseline model is defined as

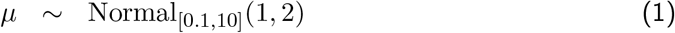

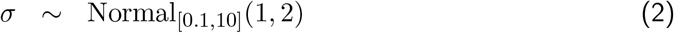

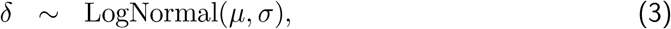

where Normal_[*l,u*]_ denotes a truncated normal distribution and the priors for *µ* and *σ* are only weakly informative and positive.

The fully hierarchical model shown in Figure 4 turned out to be infeasibly costly to compute on the large amounts of data generated during the pandemic. As a less sophisticated variant, we use a manual approximation to the hierarchical model by first fitting a global model for the delay distribution to all cases in Germany, then a model with means and variances per state of the form

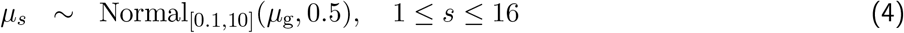

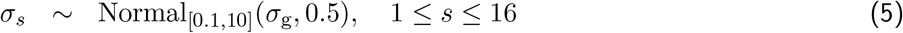

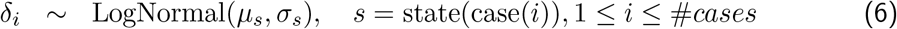

and, finally, fitting a more complex model with additional covariates (here: age group, gender, district, weekday, and number of weeks since the start of the pandemic):

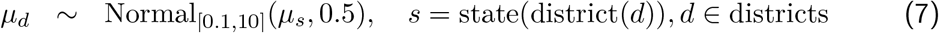

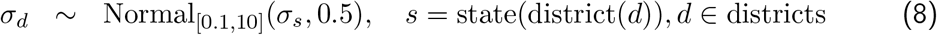

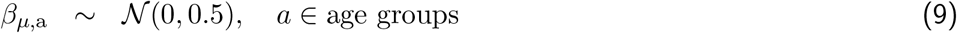

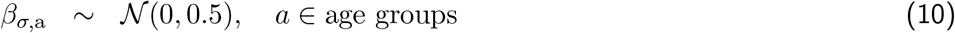

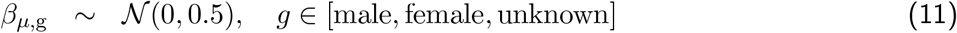

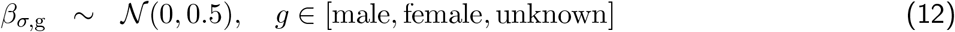

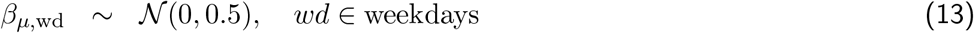

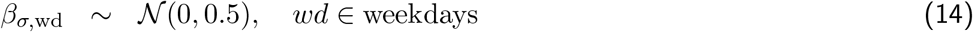

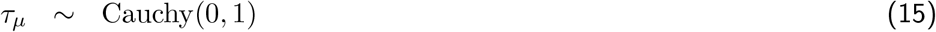

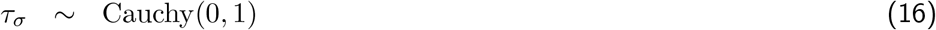

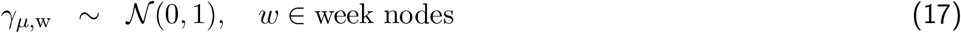

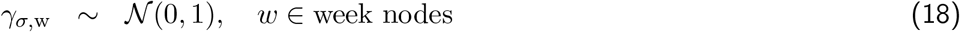

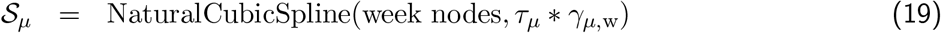

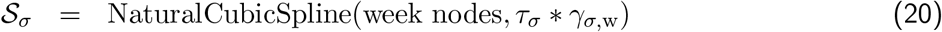

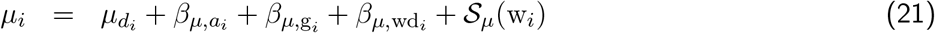

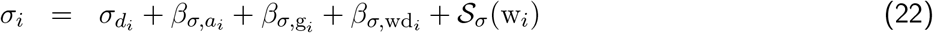

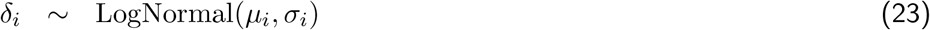

### 4.4 Nowcasting model

In this work, we use a nowcasting model based on [13], which in turn extends earlier work by [16]. These approaches use a hierarchical Bayesian model composed of two parts: a model for the delay distribution *δ* as a function of time (*P* (*δ*_*i*_ = *d*|*x*_*i*_ = *t*) := *p*_*t,d*_) and a model for the expected number of infections per day (*E*[*N* (*t*, ∞)] =: *λ*_*t*_).

For the first part, we can either plug in the delay distribution obtained during imputation (cf. Section 2.2) or fit a new model, such as the one proposed in [13], which starts from a discrete time hazard model with *h*_*t,d*_ = *P* (*δ* = *d*|*δ* ≥ *d, W*_*t,d*_) with time- and delay-dependent covariates *W*_*t,d*_ and with logit 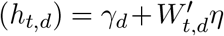, for *d* = 0,…*D* − 1, with bias *γ*_*d*_ and with *h*_*t,D*_ = 1. In this approach, the covariates *W*_*t,d*_ can represent different features of interest.

Like [13], we use a first-order spline to include a general time dependence and factor variables to represent weekdays and holidays. The probabilities *p*_*t,d*_ can be obtained from the hazard model through *p*_*t*,0_ = *h*_*t*,0_ and 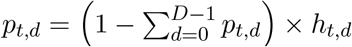.

For the second part of the hierarchical nowcasting model, we again follow [13] and use the Gaussian random walk

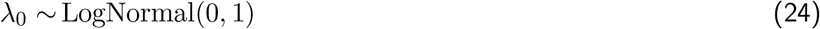

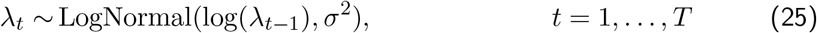

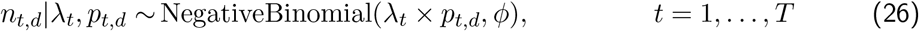

where *ϕ* is the overdispersion parameter of the negative binomial distribution. Appropriate priors for the model are again taken from [13] and the implementation referenced therein.

We finally obtain 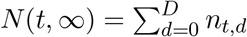.

The number of parameters of this model grows with *T* × *D*, where *T* is the day at which the nowcast is to be evaluated. Hence, the number of variables grows linearly with each day of the pandemic, rendering sampling increasingly time- and memory-consuming. To prevent this growth from turning nowcasting infeasible, we limit the amount of history we track in the Gaussian random walk, i.e., instead of setting *t* = 0 to the first day of the Covid-19 pandemic, we choose a fixed time interval of size Δ *> D* (set to the last Δ = 50 days in our implementation). Since *λ*_0_ = *E* [*N* (0, ∞)] equals the number of infections happening at day *t* = 0, we need to adapt Eq. (24) accordingly.

### 4.5 Evaluating nowcasts

Evaluating probabilistic predictions requires different scoring functions than point estimates do. For a comprehensive discussion of the topic, we refer the interested reader to [18]. Here, we briefly summarize the main evaluation measures used in this work.

Evaluation of now- and forecasts is relatively straightforward if the computational model yields the full predictive probability distribution. Given a test datum *y*, common measures are the logarithmic score [12]

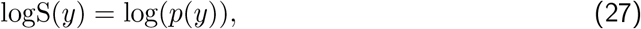

where *p*(·) denotes the probability density function used for prediction, or the continuous ranked probability score [11]

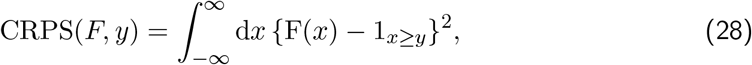

where *F* (·) denotes the cumulative distribution function used for prediction. In [13] the coverage frequency of the 95% prediction interval is used as well.

In many practical situations, the full predictive distribution is not available. In those cases, we typically have access to the predictive mean or median and, optionally, several quantiles of the cumulative distribution function. Assuming that we are given the predictive median *m* and a collection of *K* central prediction intervals (PIs) at levels (1 − *α*_1_) *<* (1 − *α*_2_) *<* · · · *<* (1 − *α*_*K*_), and weights *w*_*k*_, *k* = 1 … *K*, the weighted interval score (WIS) is defined as

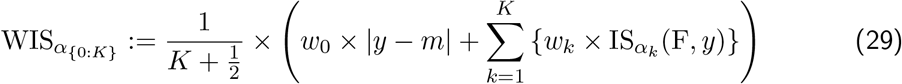

where the interval score IS_*α*_ for the central (1 − *α*) × 100% PI is defined as

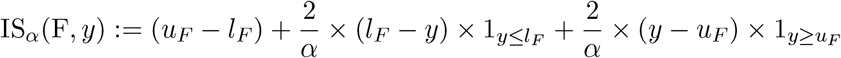

and where *l*_*F*_ and *u*_*F*_ denote the 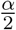-quantile and 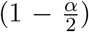-quantile of F, respectively. Following [18], we use 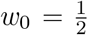 and 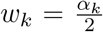 for *k* = 1 … *K*, as this choice approximates the CRPS score for large values of *K* and equidistant *α*_*k*_.

## Data Availability

All source code for the deployment and operation of the CorCast system and for the district-level models for German infection numbers will be made available upon publication.

https://github.com/hildebrandtlab/Covid19Nowcast.jl

## Authors Contributions

AKH and AH conceptualized the work. AKH, KB, DT, TK, JL and AH discussed and implemented the work, BS contributed work on parallelization and optimization. All authors contributed to the conception, writing and editing of this manuscript.

## Competing Interests

The authors declare that there is no conflict of interest.

## Data and code availability

All source code for the deployment and operation of the CorCast system and for the district-level models for German infection numbers will be made available upon publication at https://github.com/hildebrandtlab/Covid19Nowcast.jl.

## Footnotes

1 This is probably due to conform with privacy regulations.

2 Owing to Germany’s slightly archaic setup of monitoring of infectious diseases, *r*_*i*_ often differs from 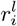, with a gap that can vary quite significantly from district to district, weekday to weekday, or week to week.

3 In practice, we replace *δ*_*i*_ by *δ*_*i*_ + 1 during inference and correct accordingly in the imputation stage. This facilitates inference, as our target distributions typically predict a probability of 0 for delays of 0. These do, however, occur in the data.

4 The details of the implementation are rather complex to handle a variety of special cases.

5 and on a more philosophical level, a different interpretation of probability. [19] provides further insights into the foundations and philosophical implications of Bayesian thinking.

## Notes

### Competing Interest Statement

The authors have declared no competing interest.

### Funding Statement

No external funding was received.

